# Ethnic minority groups in England and Wales - factors affecting the size and timing of elevated COVID-19 mortality: a retrospective cohort study linking Census and death records

**DOI:** 10.1101/2020.08.03.20167122

**Authors:** Daniel Ayoubkhani, Vahé Nafilyan, Chris White, Peter Goldblatt, Charlotte Gaughan, Louisa Blackwell, Nicky Rogers, Ami Banerjee, Kamlesh Khunti, Myer Glickman, Ben Humberstone, Ian Diamond

## Abstract

**Objectives:** To estimate population-level associations between ethnicity and coronavirus disease 2019 (COVID-19) mortality, and to investigate how ethnicity-specific mortality risk evolved over the course of the pandemic.

**Design:** Retrospective cohort study using linked administrative data.

**Setting:** England and Wales, deaths occurring 2 March to 15 May 2020.

**Participants:** Respondents to the 2011 Census of England and Wales aged ≤100 years and enumerated in private households, linked to death registrations and adjusted to account for emigration before the outcome period, who were alive on 1 March 2020 (*n*=47,872,412).

**Main outcome measure:** Death related to COVID-19, registered by 29 May 2020.

**Statistical methods:** We estimated hazard ratios (HRs) for ethnic minority groups compared with the White population using Cox regression models, controlling for geographical, demographic, socio-economic, occupational, and self-reported health factors. HRs were estimated on the full outcome period and separately for pre- and post-lockdown periods in the UK.

**Results:** In the age-adjusted models, people from all ethnic minority groups were at elevated risk of COVID-19 mortality; the HRs for Black males and females were 3.13 [95% confidence interval: 2.93 to 3.34] and 2.40 [2.20 to 2.61] respectively. However, in the fully adjusted model for females, the HRs were close to unity for all ethnic groups except Black (1.29 [1.18 to 1.42]). For males, COVID-19 mortality risk remained elevated for the Black (1.76 [1.63 to 1.90]), Bangladeshi/Pakistani (1.35 [1.21 to 1.49]) and Indian (1.30 [1.19 to 1.43]) groups. The HRs decreased after lockdown for all ethnic groups, particularly Black and Bangladeshi/Pakistani females.

**Conclusions:** Differences in COVID-19 mortality between ethnic groups were largely attenuated by geographical and socio-economic factors, although some residual differences remained. Lockdown was associated with reductions in excess mortality risk in ethnic minority populations, which has major implications for a second wave of infection or local spikes. Further research is needed to understand the causal mechanisms underpinning observed differences in COVID-19 mortality between ethnic groups.

## Introduction

Coronavirus disease 2019 (COVID-19) is a serious infectious disease originally reported in China in December 2019, caused by novel severe acute respiratory syndrome coronavirus 2 (SARS-CoV-2). More than 660,000 deaths attributable to COVID-19 have been reported worldwide, and in the UK alone above 50,000 death registrations have mentioned COVID-19 [1] and over 130,000 people have been hospitalised with the disease [2] as of 29^th^ July 2020. As the pandemic has evolved, a growing body of evidence has emerged on the clinical risk factors associated with COVID-19 hospitalisation and death, including age, sex, obesity, and certain chronic conditions including diabetes, hypertension, lung disease and cardiovascular disease [3, 4, 5]. However, evidence also suggests that socio-economic factors may be drivers of variations in COVID-19 morbidity and mortality [6]. For instance, COVID-19 related mortality in England and Wales is nearly twice the rate in the most deprived areas compared to the least deprived [7], with the highest mortality rates among working age people observed among those working in elementary occupations or social care [8].

Heterogeneity in health status between ethnic groups is well documented across multiple countries over the course of a number of decades [9]. As the 2009 A(H1N1) influenza pandemic disproportionately affected ethnic minority groups in England [10], the relationship between ethnicity and COVID-19 outcomes is of substantial interest, particularly in England and Wales where 14% of the population reported being of non-White ethnicity at the last decennial census in 2011. Existing evidence indicates that the risk of hospitalisation and death due to COVID-19 in the UK is elevated for BAME (Black, Asian and Minority Ethnic) individuals compared to the White population [11, 12, 3]. However, the magnitude of the association varies depending on the data sources used and the covariates adjusted for [13]. In addition, not enough is known about the mechanisms driving differences in the risk of COVID-19 related death across ethnic groups.

The present study makes three important contributions to research into ethnic disparities in COVID-19 outcomes. Firstly, we used microdata from the 2011 Census of England and Wales and linked death registrations to investigate, at population level, the association between ethnicity and COVID-19 related mortality risk. Secondly, we exploited a broad range of demographic and structural socio-economic measures available in the 2011 Census, often associated with inequality in health status, to investigate whether these factors mediate the relationship between ethnicity and COVID-19 mortality risk. Thirdly, we examined how the relative risk of COVID-19 mortality between White and ethnic minority groups has evolved over the course of the pandemic, focussing on the periods before and after the “lockdown” restrictions on freedom of activity were announced by the UK Government.

## Methods

### Study design and data

In this observational, retrospective cohort study, the study population included all usual residents of England and Wales who were enumerated in private households (that is, not communal establishments) at the time of the 2011 Census (27 March 2011) and who were not known to have died before 2 March 2020. We excluded from the study population individuals who entered the UK in the year before the Census took place due to their high propensity to have left the UK prior to the study period, and those over 100 years of age at the time of the Census even if their death was not linked. Our study population therefore consisted of 47,872,412 individuals aged 9 years or over at 2 March 2020 (the youngest possible, given the nine-year lag from enumeration of the study population at the 2011 Census).

We used a unique, newly-created linked dataset that combines 2011 Census records and death registrations for England and Wales, created by linking the 2011 Census to NHS Patient Register records between 2011 and 2013, with a linkage rate of 94.6% (for further details, see the Supplementary Appendix on Census to Patient Register linkage rates by characteristics of interest).

Deaths were linked to the 2011 Census using NHS Number, with 89.9% of deaths occurring between 27 March 2011 and 1 March 2020 being linked to the 2011 Census. We linked deaths occurring between 2 March 2020 and 15 May 2020 that were registered by 29 May 2020 using NHS Number and a deterministic match key linkage method where NHS Number was unavailable, achieving a linkage rate of 90.8% of deaths. The unmatched deaths comprise people not present in the UK at the 2011 Census, people who arrived in the UK in the year before the Census (and were excluded from the study), and people who were present at Census but not enumerated.

The study dataset did not contain any information on whether individuals have left the country. To avoid biasing our denominators, we derived and applied weights reflecting the probability to have remained in the country between March 2011 and March 2020, based on data from the NHS Patient Register and the International Passenger Survey (IPS). For further information, see the Supplementary Appendix on the methodology for adjusting for emigration.

Despite being in the population at risk of COVID-19 death in March 2020, we did not replenish the sample with post-2011 births or immigrants; while the latter group could have been identified and in principle linked to our data, neither group are captured in the 2011 Census and therefore they have no ethnicity or covariate data recorded. Additionally, the younger population have been the least affected with COVID-19 related hospitalisation and mortality. For the same reason, individuals not enumerated at the 2011 Census (estimated to be 6.1% of the population of England and Wales [14]) were not included in our study population.

### Outcome and exposure

In this study, the outcome of interest was death involving COVID-19 that occurred between 2 March 2020 and 15 May 2020, registered by 29 May 2020. Deaths involving COVID-19 included those with an underlying cause or any mention on the death certificate of the ICD-10 codes U07.1 (COVID-19, virus identified) or U07.2 (COVID-19, virus not identified). Time-at-risk started on 2 March 2020 and ran until 15 May 2020 or date of death, whichever was earlier.

Our exposure of interest was self-reported ethnic affiliation, as stated by respondents to the 2011 Census. Ethnic breakdown was based on the Census ethnicity question, with some groups collapsed to ensure outcome counts were large enough to permit reliable estimates of hazard ratios (see Supplementary Table for more detail on how the groups were aggregated). The White ethnic group was used as the reference category in our modelling analysis as it is the largest group. Ethnicity was imputed in 3.0% of 2011 Census returns due to item non-response, using the imputation methodology employed by the Office for National Statistics (ONS) across all 2011 Census variables [15].

### Covariates

We controlled for demographic and structural socio-economic factors that may be associated with the risk of COVID-19 mortality (Table 1), through either the propensity to become infected or the propensity to die once infected (which could not be distinguished using the data available for this study). Such factors are likely to confound and/or mediate the relationship between ethnicity and COVID-19 mortality risk.

**Table 1.**
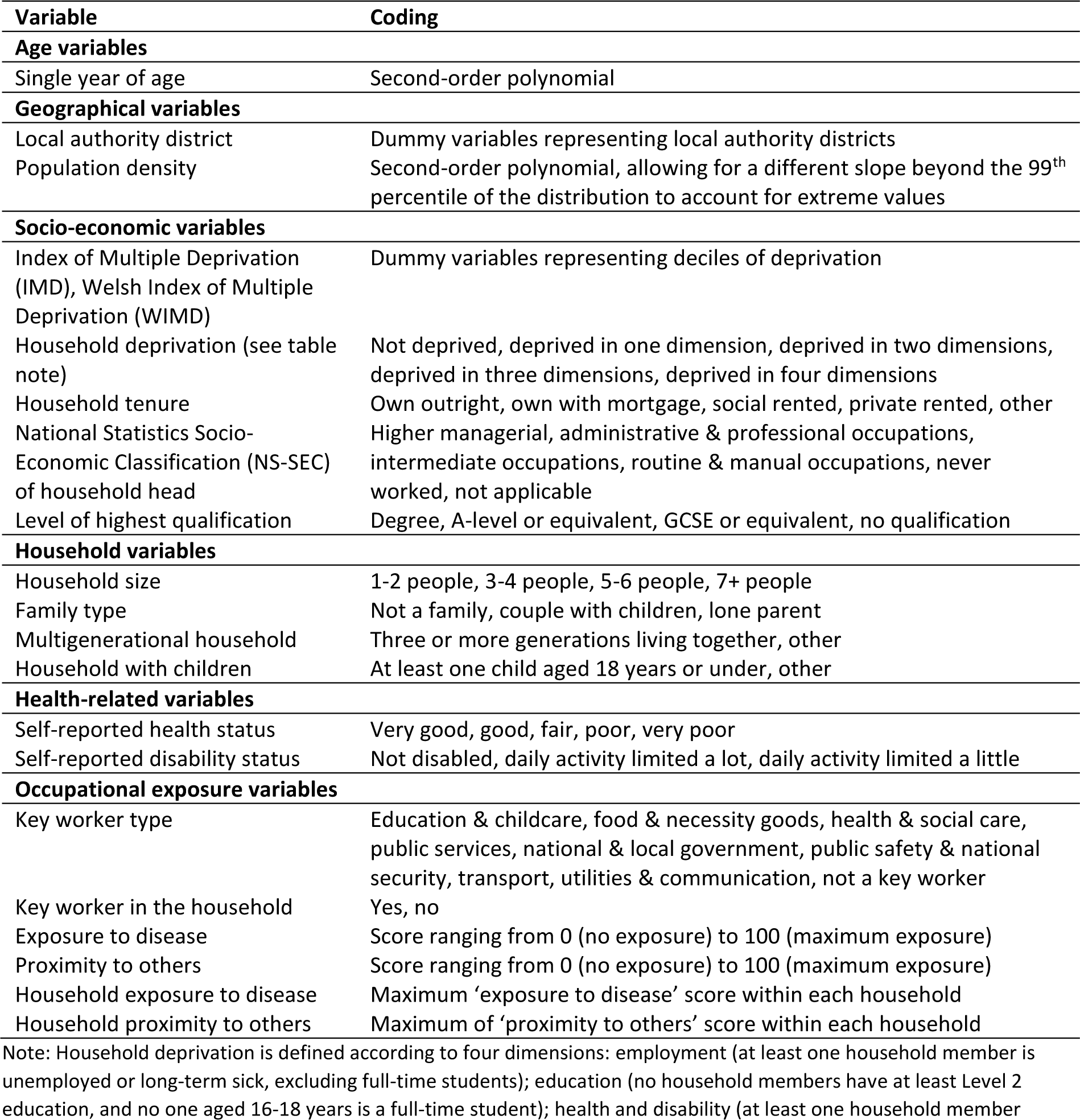
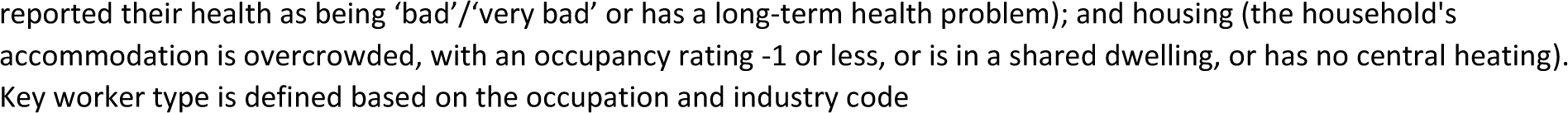
Covariates included in the Cox regression models

| Variable | Coding |
| --- | --- |
| **Age variables** |  |
| Single year of age | Second-order polynomial |
| **Geographical variables** |  |
| Local authority district | Dummy variables representing local authority districts |
| Population density | Second-order polynomial, allowing for a different slope beyond the 99<sup>th</sup> percentile of the distribution to account for extreme values |
| **Socio-economic variables** |  |
| Index of Multiple Deprivation (IMD), Welsh Index of Multiple Deprivation (WIMD) | Dummy variables representing deciles of deprivation |
| Household deprivation (see table note) | Not deprived, deprived in one dimension, deprived in two dimensions, deprived in three dimensions, deprived in four dimensions |
| Household tenure | Own outright, own with mortgage, social rented, private rented, other |
| National Statistics Socio-Economic Classification (NS-SEC) of household head | Higher managerial, administrative & professional occupations, intermediate occupations, routine & manual occupations, never worked, not applicable |
| Level of highest qualification | Degree, A-level or equivalent, GCSE or equivalent, no qualification |
| **Household variables** |  |
| Household size | 1-2 people, 3-4 people, 5-6 people, 7+ people |
| Family type | Not a family, couple with children, lone parent |
| Multigenerational household | Three or more generations living together, other |
| Household with children | At least one child aged 18 years or under, other |
| **Health-related variables** |  |
| Self-reported health status | Very good, good, fair, poor, very poor |
| Self-reported disability status | Not disabled, daily activity limited a lot, daily activity limited a little |
| **Occupational exposure variables** |  |
| Key worker type | Education & childcare, food & necessity goods, health & social care, public services, national & local government, public safety & national security, transport, utilities & communication, not a key worker |
| Key worker in the household | Yes, no |
| Exposure to disease | Score ranging from 0 (no exposure) to 100 (maximum exposure) |
| Proximity to others | Score ranging from 0 (no exposure) to 100 (maximum exposure) |
| Household exposure to disease | Maximum 'exposure to disease' score within each household |
| Household proximity to others | Maximum of 'proximity to others' score within each household |

Note: Household deprivation is defined according to four dimensions: employment (at least one household member is unemployed or long-term sick, excluding full-time students); education (no household members have at least Level 2 education, and no one aged 16-18 years is a full-time student); health and disability (at least one household member
reported their health as being 'bad'/'very bad' or has a long-term health problem); and housing (the household's accommodation is overcrowded, with an occupancy rating -1 or less, or is in a shared dwelling, or has no central heating). Key worker type is defined based on the occupation and industry code

All models were adjusted for single year of age, included as a second-order polynomial to account for the non-linear relationship between age and the hazard of death involving COVID-19. Geographical variables included the local authority district of residence at the time of the 2011 Census and official estimates of mid-2018 population density for the Lower Super Output Area (LSOA) of residence.

To account for neighbourhood deprivation, we adjusted for the decile of the Index of Multiple Deprivation (IMD) 2015 and Welsh Index of Multiple Deprivation (WIMD) 2014 of the LSOA of residence at the time of the Census. The IMD and WIMD are overall measures of deprivation in the local area based on factors such as income, employment and health [16, 17].

As further measures of socio-economic position, we used the level of household deprivation, a summary measure of disadvantage based on four selected household characteristics (employment, education, health and housing), the tenure of the household, and the National Statistics Socio-economic Classification (NS-SEC) of the household reference person. The NS-SEC is not defined for respondents aged 75 or over at the time of the Census, so we included individuals’ highest level of qualification as an additional indicator of socio-economic position.

Our measures of household composition and circumstances included the number of people in the household, the family type, and binary variables for living in a multigenerational household (defined as three generations living together) or with any children (aged 18 years or under). We included self-reported health status and a variable indicating whether the individual was activity restricted (disabled) at the 2011 Census.

Finally, in the fully adjusted model we incorporated measures of occupational exposure as recorded at the 2011 Census. We included a variable indicating if an individual was a key worker, and if so, what type. We also included a binary variable indicating if anyone in the household was a key worker. We accounted for exposure to diseases and contact with others using scores ranging from 0 (no exposure) to 100 (maximum exposure), derived using O*NET data based on US Standard Occupational Classification (SOC) codes mapped to UK SOC codes [18]. We assigned these scores based on individuals’ occupations and then derived the maximum value among all household members.

### Statistical Analyses

#### Age-standardised mortality rates

To examine how the absolute risk of COVID-19 related death varied across ethnic groups, we calculated age-standardised mortality rates for the different groups. These can be interpreted as deaths per 100,000 of the population during the analysis period, where the age-by-sex distribution within each ethnic group was standardised to the overall distribution in the study population (after applying the emigration weights to the population denominators).

#### Cox proportional hazard models

We used Cox proportional hazard models to assess whether differences in the risk of COVID-19-related death across ethnic groups could be accounted for by the factors listed in Table 1. When fitting the Cox models, we included all individuals who died during the analysis period and a weighted 1% random sample of those who did not.

We estimated separate models for males and females, as the risk of death involving COVID19 differs markedly by sex [3]. We estimated models that only adjust for age before adding groups of control variables, as defined above, step by step and assessing how these affected the estimated hazard ratios between ethnic groups. In sensitivity analysis, we tested whether interacting each of the controls with a binary variable indicating if the individual was aged 70 years or over and a variable indicating whether the individual lived in London had any effect on the hazard ratios for ethnic minority groups.

#### Assessing the proportional hazards assumption

We used Schoenfeld residuals from the fitted Cox models, smoothed using generalised additive models, to assess whether relative differences in the hazard of COVID-19 mortality between the ethnic groups changed over the course of the pandemic [19]. This provided a time-varying estimate of the association between belonging to an ethnic minority group and the log hazard ratio of a COVID-19 related death, conditional on the covariates included in the model.

To address potential non-proportionality in hazards between the White and ethnic minority groups, we extended the Cox models by allowing for time-dependent ethnicity coefficients. We selected 23 March 2020 (the date on which legally enforceable restrictions to freedom of activity were announced by the UK Government) plus three weeks (to allow for a lag between lockdown coming into force and its likely impact on COVID-19 mortality rates) as a landmark date and split the follow-up time of people who were still in the study after this date into pre- and post-lockdown periods, with pre-lockdown outcomes recorded as censored. We fitted Cox models with stratification of the ethnicity estimates on pre-/post-lockdown period, thus assuming a step change in the hazard ratios at lockdown date plus three weeks. In sensitivity analysis, we investigated the impact on the estimated hazard ratios of varying the 21-day lag duration by ±7 days.

All statistical analyses were conducted using R version 3.5 [20].

#### Public involvement

The ONS received approximately 2,000 responses from members of the public and other interested parties to the 2011 Census Consultation [21], which informed the content of the Census questionnaire.

### Results

#### Characteristics of the study population

In our study population of 47,872,412 usual residents in England and Wales in 2011 who were still alive on 2 March 2020, just over half (51.6%) were female, the mean age was 47 years (minimum 9 years, maximum 110 years) and 86.4% reported being from a White ethnic background (Table 2). Over the outcome period (2 March 2020 to 15 May 2020), mean follow-up time was 73.9 days (standard deviation 2.2 days) and 37,956 individuals (0.08%) died of COVID-19.

**Table 2.** Distributions of study variables, stratified by White/non-White ethnicity

| Variable | Level | All people | White | Non-White | Cohen's *d* |
| --- | --- | --- | --- | --- | --- |
| Age (years) | Mean | 46.5 | 47.8 | 38.1 | 0.45 |
| Sex | Male | 48.4% | 48.5% | 48.4% | 0.00 |
|  | Female | 51.6% | 51.5% | 51.6% | 0.00 |
| Ethnicity | White | 86.4% | - | - | - |
|  | Bangladeshi and Pakistani | 3.0% | - | - | - |
|  | Black | 3.2% | - | - | - |
|  | Chinese | 0.6% | - | - | - |
|  | Indian | 2.6% | - | - | - |
|  | Mixed | 2.1% | - | - | - |
|  | Other | 2.2% | - | - | - |
| Population density (people per square kilometre) | Mean | 4,424 | 3,829 | 8,211 | 1.00 |
| National Statistics Socio-Economic Classification (NS-SEC) of household head | Higher managerial, administrative and professional occupations | 30.7% | 31.4% | 26.2% | 0.11 |
|  | Intermediate occupations | 18.7% | 18.6% | 19.2% | 0.01 |
|  | Routine and manual occupations | 25.9% | 26.0% | 25.4% | 0.01 |
|  | Never worked, unemployed, NEC | 2.9% | 2.4% | 6.5% | 0.25 |
|  | Not applicable | 21.8% | 21.6% | 22.7% | 0.03 |
| Tenure of household | Owned outright | 24.3% | 25.7% | 15.4% | 0.24 |
|  | Owned with a mortgage | 42.6% | 43.4% | 37.1% | 0.13 |
|  | Shared ownership | 0.7% | 0.7% | 0.8% | 0.02 |
|  | Social rented from council | 8.2% | 7.6% | 12.0% | 0.16 |
|  | Other social rented | 7.1% | 6.6% | 10.2% | 0.14 |
|  | Private rented | 16.2% | 15.1% | 23.0% | 0.21 |
|  | Living rent free | 1.0% | 0.9% | 1.5% | 0.06 |
| Level of highest qualification | No qualification | 15.4% | 16.0% | 11.9% | 0.11 |
|  | Level 1: 1-4 GCSE/O-Level | 11.2% | 11.7% | 8.5% | 0.10 |
|  | Level 2: 5+ GCSE/O levels | 12.8% | 13.5% | 8.8% | 0.14 |
|  | Apprenticeship | 2.9% | 3.2% | 0.7% | 0.15 |
|  | Level 3: 2+ A Levels or equivalent | 10.1% | 10.5% | 7.5% | 0.10 |
|  | Level 4+: degree or above | 22.9% | 22.8% | 23.9% | 0.03 |
|  | Other | 4.4% | 3.7% | 8.8% | 0.24 |
|  | No code required | 20.2% | 18.6% | 29.9% | 0.28 |
| Index of Multiple Deprivation (IMD), Welsh Index of Multiple Deprivation (WIMD) | Decile 1 | 9.7% | 8.2% | 19.2% | 0.38 |
|  | Decile 2 | 9.8% | 8.5% | 18.1% | 0.33 |
|  | Decile 3 | 9.9% | 9.2% | 14.8% | 0.19 |
|  | Decile 4 | 9.9% | 9.7% | 11.4% | 0.06 |
|  | Decile 5 | 10.0% | 10.1% | 9.3% | 0.03 |
|  | Decile 6 | 10.1% | 10.5% | 7.3% | 0.11 |
|  | Decile 7 | 10.1% | 10.8% | 5.7% | 0.17 |
|  | Decile 8 | 10.1% | 10.9% | 5.1% | 0.19 |
|  | Decile 9 | 10.2% | 11.1% | 4.6% | 0.21 |
|  | Decile 10 | 10.2% | 11.1% | 4.5% | 0.22 |
| Multigenerational household | Yes | 5.5% | 5.0% | 9.1% | 0.18 |
|  | No | 94.5% | 95.0% | 90.9% | 0.18 |
| Child in household | Yes | 49.8% | 47.0% | 67.2% | 0.41 |
|  | No | 50.2% | 53.0% | 32.8% | 0.41 |
| Household size | 1-2 people | 39.7% | 42.4% | 22.4% | 0.41 |
|  | 3-4 people | 44.2% | 44.4% | 42.5% | 0.04 |
|  | 5-6 people | 14.1% | 12.0% | 27.2% | 0.44 |
|  | 7+ people | 2.1% | 1.1% | 7.9% | 0.48 |
| Family status | Not in a family | 15.6% | 15.7% | 14.7% | 0.03 |
|  | In a couple family | 71.0% | 71.7% | 66.5% | 0.11 |
|  | In a lone parent family | 13.5% | 12.6% | 18.8% | 0.18 |
| Keyworker type | Not key worker | 84.4% | 83.8% | 88.0% | 0.11 |
|  | Education & childcare | 4.6% | 4.9% | 2.5% | 0.11 |
|  | Food & necessary goods | 0.8% | 0.8% | 0.5% | 0.02 |
|  | Health & social care | 5.5% | 5.5% | 5.8% | 0.02 |
|  | Key public services | 1.2% | 1.2% | 0.8% | 0.02 |
|  | National & local government | 0.6% | 0.6% | 0.4% | 0.03 |
|  | Public safety & national security | 1.2% | 1.3% | 0.5% | 0.08 |
|  | Transport | 0.9% | 0.9% | 0.6% | 0.04 |
|  | Utilities & communication | 1.0% | 1.0% | 0.9% | 0.02 |
| Key worker in household | Yes | 15.9% | 16.5% | 12.3% | 0.06 |
|  | No | 84.1% | 83.8% | 87.7% | 0.06 |
| Exposure to disease score (0-100) | Mean (individual) | 14.7 | 15.0 | 12.8 | 0.11 |
|  | Mean (household) | 28.7 | 28.4 | 30.5 | 0.09 |
| Proximity to others score (0-100) | Mean (individual) | 45.4 | 46.9 | 35.5 | 0.38 |
|  | Mean (household) | 67.8 | 67.9 | 67.1 | 0.05 |
| Self-reported health status | Very good | 49.7% | 49.2% | 53.3% | 0.08 |
|  | Good | 35.0% | 35.2% | 33.6% | 0.03 |
|  | Fair | 11.5% | 11.7% | 9.7% | 0.06 |
|  | Poor | 3.1% | 3.1% | 2.8% | 0.02 |
|  | Very poor | 0.7% | 0.7% | 0.8% | 0.01 |
| Long-term health problem or disability | Daily activities not limited | 86.1% | 85.6% | 89.4% | 0.11 |
|  | Daily activities limited a little | 5.6% | 5.8% | 4.6% | 0.05 |
|  | Daily activities limited a lot | 8.3% | 8.7% | 5.9% | 0.10 |

Note: Study population consists of usual residents in England and Wales in 2011 who were still alive on 2 March 2020, down weighted to account for emigration over this period. Summary statistics calculated on all individuals who died and a 1% random sample of those who did not die, weighted to represent the full study population. Cohen's $d$ is a measure of effect size between the White and non-White populations. Values of $d=0.2$ , $d=0.5$ and $d=0.8$ are indicative of "small", "medium" and "large" effect sizes, respectively [22].

Compared with the White population, individuals in the non-White group: had a lower mean age; tended to reside in more densely populated areas; were more likely to have never worked or be unemployed; were less likely to own their own home and more likely to rent; were more likely to live in a deprived neighbourhood; and were more likely to live in a larger household, with at least one household member being a child.

#### Risk of COVID-19 mortality by ethnicity

For both males and females, age-standardised mortality rates (ASMRs) of COVID-19 related death (see Supplementary Table 2) were greatest among individuals identifying as Black (250.6 and 116.9 deaths per 100,000 people, respectively) and lowest among those identifying as White (87.3 and 51.8 deaths per 100,000 people, respectively). The ASMRs indicate greater levels of absolute risk among all ethnic minority groups compared with the White population.

In terms of age-adjusted hazard ratios, males and females from all ethnic minority groups (except females of Chinese ethnicity) were at greater risk of death involving COVID-19 compared with those of White ethnicity (Figure 1, Model 1). The rate of death involving COVID-19 adjusted for age was 3.13 [95% confidence interval (CI): 2.93-3.34] times greater for Black males than White males, and 2.40 [2.20-2.61] times greater for Black females than White females. People of Bangladeshi/Pakistani, Indian, Mixed and Other ethnic backgrounds also had raised rates of death involving COVID-19 compared with those of White ethnicity.

**Figure 1.**
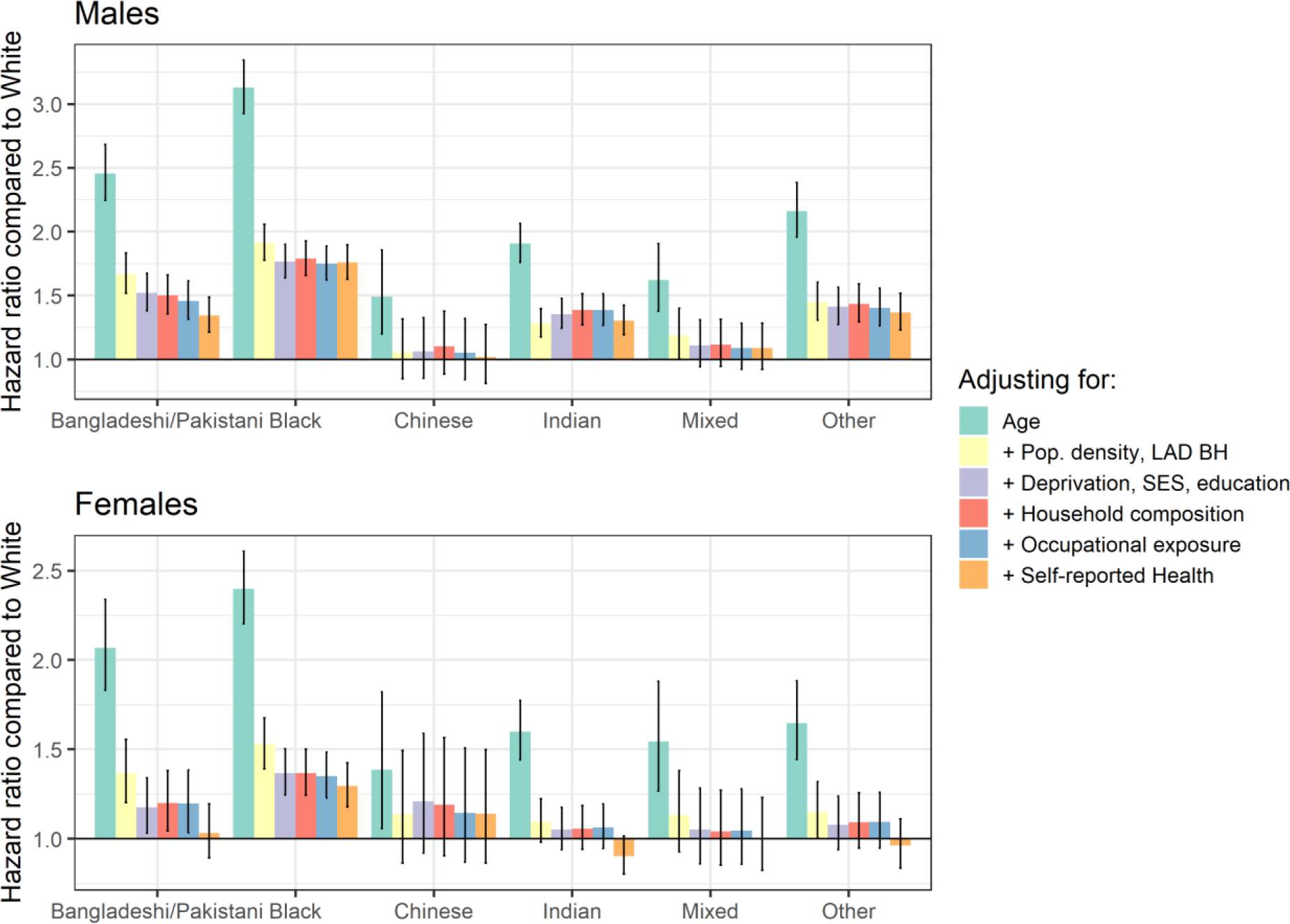
Hazard ratios for COVID-19 related death for ethnic minority groups compared to the White population, stratified by sex Note: Results obtained from Cox regression models adjusted for age, population density, area and household deprivation, Socio-economic Status (SES), household composition, occupational exposure, self-reported health, with baseline hazards specific to local authority district. Errors bars represent limits of 95% confidence intervals of the hazard ratio. LAD BH denotes baseline hazard specific for each local authority district. Details of the covariates can be found in Table 1. Numerical results can be found in Supplementary Tables 3 and 4.

Including baseline hazards that are specific to each local authority district and controlling for population density substantially reduced the estimated differences in the rate of death involving COVID-19 across ethnic groups (Model 2). The attenuating effect of controlling for geography was greatest for Chinese males and Indian females, for whom the excess risk relative to the White group decreased by 89% and 84% respectively, relative to the age-adjusted model.

Further adjusting for deprivation and socio-economic position (Model 3), household composition (Model 4) and occupational exposure (Model 5) had a small impact on the hazard ratios, but not negligible for people of Bangladeshi and Pakistani background and Black males. Controlling for health and disability status (Model 6) at the time of the 2011 Census reduced the estimated hazard ratios for some groups, especially individuals of Bangladeshi/Pakistani or Indian ethnicity.

Taken together, the characteristics in the fully adjusted model for females statistically explained the differences in risk compared to the White population for all ethnic minority groups except those of Black ethnicity, as their hazard ratio remained at 1.29 [1.17-1.42] after adjustment.

Adjusting for these characteristics also reduced the estimated hazard ratios for males, but they remained larger than one for all groups other than Chinese and Mixed ethnic backgrounds. Compared to White males, the rate of death involving COVID-19 was 1.77 [1.64-1.91] times greater for Black males, 1.35 [1.22-1.50] times greater for Bangladeshi and Pakistani males, and 1.31 [1.19-1.43] times greater for Indian males.

#### Risk of COVID-19 mortality before and after lockdown

The smoothed Schoenfeld residuals by ethnic group (see Supplementary Supplementary Figure and 2) indicated a non-constant hazard ratio of deaths involving COVID-19 over the course of the pandemic for some groups, with the hazard ratios for these groups decreasing with time. For the Bangladeshi/Pakistani and Chinese groups for males and the Black group for females, the log hazard ratios tended to zero midway through the outcome period, indicating diminished differences in risk between ethnic minority groups and the White population.

**Figure 2.**
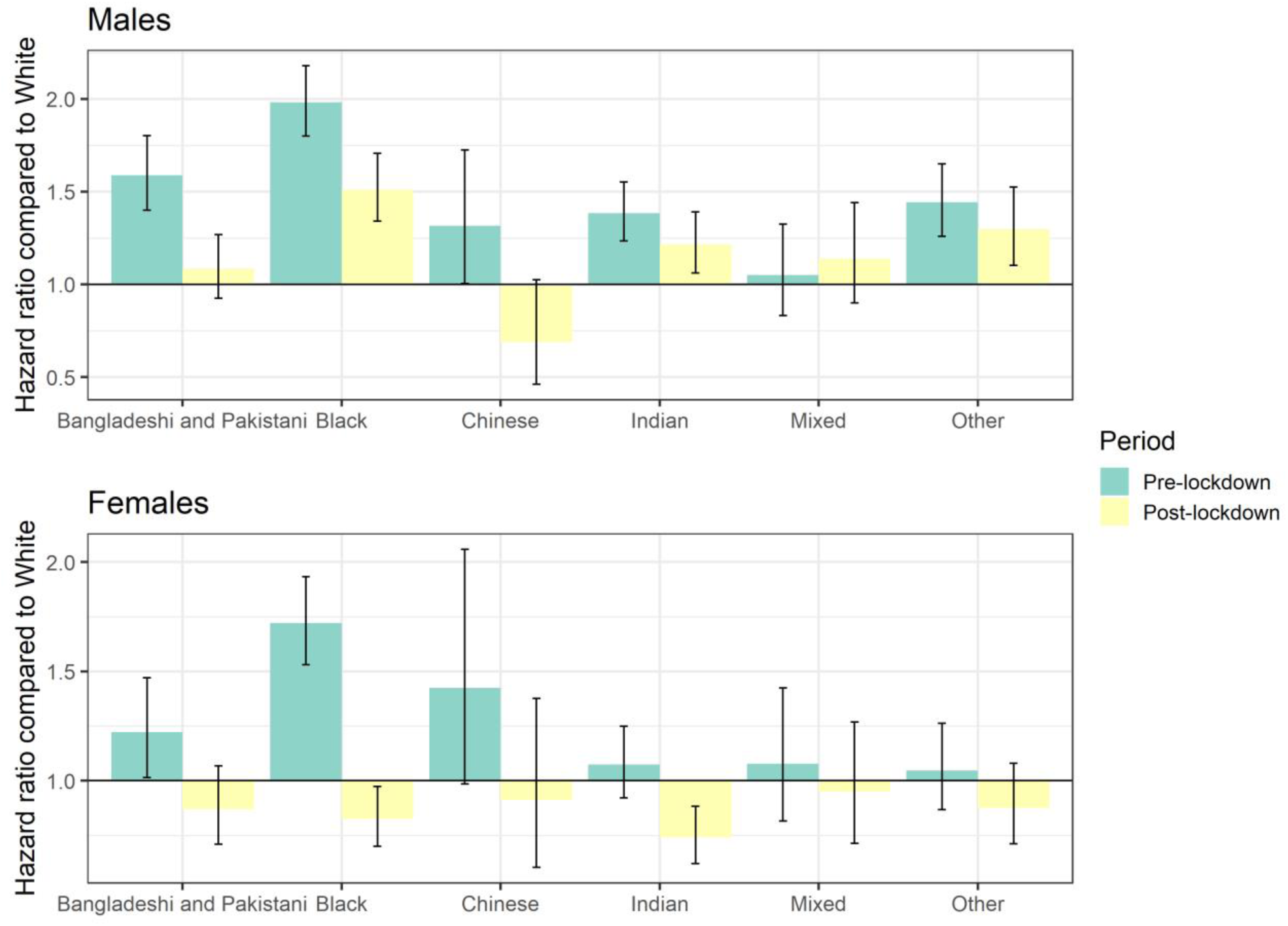
Fully adjusted hazard ratios for COVID-19 related death for ethnic minority groups compared to the White population, before and after lockdown plus 21 days, stratified by sex Note: Results obtained from a Cox regression model adjusted for age, population density, deprivation, SES, household composition, occupational exposure, self-reported health, with baseline hazards specific to local authority district. Errors bars represent limits of 95% confidence intervals of the hazard ratio. Estimates for each ethnic group are stratified by a time variable indicating pre- and post-lockdown periods.

The ASMRs decreased after lockdown for males of all ethnic minority groups, most notably by 61.8 deaths per 100,000 of the Black population, but increased by 12.1 deaths per 100,000 of the White population (Supplementary Table 5). Similar patterns were observed for females, with ASMRs after lockdown decreasing by 42.4 deaths per 100,000 of the Black population but increasing by 13.3 deaths per 100,000 of the White population. The increase in ASMRs for the White population may be due to deaths in care homes peaking later than throughout the rest of the community (See Supplementary Figure 3). The White population was on older average than other ethnic groups (Table 2), and therefore more likely to live in care settings.

When modelling the effect of lockdown in our fully adjusted Cox regression models, all the estimated hazard ratios decreased after the lockdown for both males and females (Figure 2). This finding is consistent with the results from the smoothed Schoenfeld residuals and indicates the elevated risk of COVID-19 mortality among ethnic minority groups relative to the White population was more pronounced before the lockdown restrictions were announced.

For females, the rate of mortality involving COVID-19 was elevated for the Bangladeshi/Pakistani and Black groups compared with the White population over the pre-lockdown period, even after accounting for individual and household characteristics, with hazard ratios of 1.22 [1.01-1.47] and 1.72 [1.53-1.93] respectively. After the lockdown, the hazard ratios decreased to 0.87 [0.71-1.07] for the Bangladeshi/Pakistani population and 0.83 [0.70-0.97] for the Black population.

The adjusted differences between pre- and post-lockdown hazard ratios were generally less pronounced for males than females. The Black, Indian and Other ethnic minority groups continued to experience a greater rate of COVID-19 mortality than the White population, but with reduced hazard ratios of 1.18 [1.04-1.34], 1.51 [1.34-1.71], 1.22 [1.06-1.39] and 1.30 [1.10-1.52] respectively.

Results obtained using shorter and longer lag durations for capturing post-lockdown deaths (14 and 28 days instead of 21 days) were similar to those in the main analysis, although less precisely estimated when using a lag of 28 days (see Supplementary Figures 4 and 5).

## Discussion

### Principal findings

Our paper has three novel findings. Firstly, using population-level data we observed a substantially elevated risk of COVID-19 related death among ethnic minority groups. Secondly, this elevated risk was largely – but not entirely – mediated by location, living circumstances, socio-economic factors, occupational exposure, and self-reported health status. Thirdly, differences in COVID-19 mortality rates between ethnic groups were considerably reduced following the introduction of lockdown measures in the UK.

### Comparison with related studies

Our finding that ethnic minority groups were at greater risk of COVID-19 related death compared with the White population is broadly consistent with other UK-based research published to date. An analysis of laboratory confirmed cases of COVID-19 in England found that some, but not all, ethnic minority groups were at greater risk of death after adjusting for demographics and deprivation, with Black and Asian (particularly Bangladeshi) groups having the highest mortality rates [12]. However, the tested sample is unlikely to be representative of the general population, which could bias the estimates of the relationship between ethnicity and COVID-19 mortality [23]. A retrospective study of electronic primary care records covering approximately 40% of patients in England found elevated COVID-19 mortality risk for Black and South Asian individuals after controlling for deprivation and clinical risk factors [3]. Despite its relatively large size and richness of clinical information, the data may not be fully representative of the full population of England and do not include socio-economic covariates other than area deprivation.

Studies based on COVID-19 surveillance data suggest that Black and South Asian individuals were more likely than White British people to test positive for COVID-19 after controlling for demographic, socio-economic, behavioural and health-related covariates [24, 25]. These findings shed some light on our own results, as observed inequality in the risk of dying with COVID-19 may partly be a result of heterogeneity in the risk of infection. Our results could also be driven by differences in prognosis. Studies examining differences in outcomes among hospitalised patients found that patients from ethnic minority groups were more likely to be admitted to critical care [26, 11] and had higher in-hospital mortality rates among COVID-19 cases in England [11].

Beyond the UK, a systematic review of global literature on the association between ethnicity and COVID-19 infection and outcomes concluded that BAME individuals were likely to be at elevated risk [13]. However, published evidence available at the time of the review was found to be limited, with much of the emerging data originating from grey literature and preprint articles.

Whilst there is evidence that lockdown measures reduced infection [27], little is known on whether these measures affected ethnic groups differently.

### Potential mechanisms

We found that area of residence and population density together accounted for over half of the excess hazard of COVID-19 mortality across all ethnic minority groups. These groups tend to be concentrated in large urban deprived areas in the UK, which have been more severely affected by the COVID-19 pandemic in terms of both infection [28] and mortality rates [7].

The elevated risk of COVID-19 mortality for ethnic minority groups compared to the White population was also statistically attenuated by the inclusion of socio-economic variables in our models, suggesting that these factors collectively mediate much of the observed risk. The existence of a social gradient in general health status and life expectancy is long-established [29], and recent official data have shown that White individuals in England are less likely to live in deprived neighbourhoods than non-White individuals [30]. These findings emphasise the importance of policy interventions aimed at reducing socio-economic inequalities as a means of reducing heterogeneity in COVID-19 (and broader) mortality risk, particularly among ethnic minority groups who are observed to accrue smaller gains from socio-economic improvements than the White population [31], and highlights the need to study the health of ethnic minority groups within the context of their broader socioeconomic disadvantage.

After full adjustment we found no difference in COVID-19 mortality for females from all ethnic minority groups except those of Black ethnicity, but elevated risks remained for males from most ethnic minority groups. The lockdown measures attenuated the disparity in COVID-19 mortality across ethnic groups, suggesting that the differences are largely due to an increased risk of infection rather than a worse prognosis. However, males from most ethnic minority groups were still at higher risk of COVID-19 mortality.

The residual differences may in part be due to unobserved factors, including pre-existing conditions and other clinical risk factors not fully captured by self-reported health status. People from Black and Asian backgrounds tend to have a higher prevalence of obesity and associated conditions of diabetes, hypertension and cardiovascular disease compared to the White population [32]. These conditions are known risk factors for adverse COVID-19 outcomes and could mediate the observed differences in mortality rates between ethnic groups; however, they may also be partly attributable to the same socio-economic inequalities already captured in our models, or to specific features of the disadvantage suffered by ethnic minority populations such as structural discrimination.

Socio-economic inequality is a complex, multi-faceted, partly ecological concept. The statistical measures of socio-economic status and deprivation included in our models are unlikely to capture fully this conceptual complexity [33], nor the fact that disadvantage accumulated over the life-course may be partly responsible for observed health inequalities [34]. Given these measurement issues, we cannot rule out the possibility of structural socio-economic factors explaining more of the heterogeneity in outcomes than estimated in our study, rather than the unexplained variation necessarily being attributable to other influences.

### Strengths and limitations

The primary strength of our study lies in the use of a unique, newly linked population-level dataset based on the 2011 Census, comprising a whole population level cohort and a broad range of demographic, socio-economic and self-reported health variables. We report results disaggregated by groupings of self-reported ethnic affiliation, across the full range of ethnicities. To our knowledge, our study is the first to demonstrate an association between COVID-19 lockdown measures and ethnicity-specific mortality.

The main limitation of our study dataset is the nine-year lag between Census day and the start of the pandemic. The covariates included in our models reflect the socio-economic situations of individuals as they were in 2011, not necessarily those at the start of the COVID-19 pandemic. In addition, the study population did not include people who immigrated or were born between 2011 and 2020, and therefore did not fully represent the population at risk.

## Conclusions

Whilst inequality in COVID-19 mortality between ethnic groups was appreciably accounted for by differences in geographical and measured socio-economic factors, some residual differences in risk remained, particularly for males and during the pre-lockdown period. Further research is urgently needed to understand the causal mechanisms underpinning observed differences in COVID-19 mortality risk between ethnic groups, and the extent to which these reflect aspects of disadvantage potentially amenable to policy interventions that target the wider social determinants of health.

Lockdown measures were associated with substantial reductions in excess mortality risk in ethnic minority populations, possibly due to behavioural change throughout society and shielding from the virus among the most vulnerable. This finding has major implications in the event of a second wave of infection or local spikes in incidence, namely that placing restrictions on freedom of movement and activity may be seen as a “leveller” in terms of COVID-19 mortality rates between ethnic groups.

## Data Availability

Under the provisions of the Statistics and Registration Service Act 2007, the linked 2011 Census data used in this study are not permitted to be shared.

## Footnotes

## Acknowledgements

The authors would like to thank Karen Tingay and Neil Bannister at the Office for National Statistics, Miqdad Asaria at the London School of Economics and Nazrul Islam at the Nuffield Department of Population Health for their valuable input to the manuscript. The authors also thank Merilynn Pratt, Alexander Cooke, David Tabor and Jessica Walkeden at the Office for National Statistics for their work in preparing the study data. KK is supported by the National Institute for Health Research (NIHR) Applied Research Collaboration East Midlands (ARC EM) and the NIHR Leicester Biomedical Research Centre (BRC).

## Contributors

DA, VN, CW, CG, MG and BH contributed to the study conceptualisation and design. LB and NR contributed to preparation of the study data, and DA, VN, CW and CG performed the statistical analyses. All authors contributed to interpretation of the results. DA, VN, CW, LB and NR drafted the manuscript. PG, AB, KK, MG, BH and ID contributed to the critical revision of the manuscript. All authors approved the final manuscript. VN is the guarantor for the study. The corresponding author attests that all listed authors meet authorship criteria and that no others meeting the criteria have been omitted.

## Copyright/license for publication

The Corresponding Author has the right to grant on behalf of all authors and does grant on behalf of all authors, a worldwide licence to the Publishers and its licensees in perpetuity, in all forms, formats and media (whether known now or created in the future), to i) publish, reproduce, distribute, display and store the Contribution, ii) translate the Contribution into other languages, create adaptations, reprints, include within collections and create summaries, extracts and/or, abstracts of the Contribution, iii) create any other derivative work(s) based on the Contribution, iv) to exploit all subsidiary rights in the Contribution, v) the inclusion of electronic links from the Contribution to third party material where-ever it may be located; and, vi) licence any third party to do any or all of the above.

## Dissemination declaration

The direct dissemination to study participants is not possible because of the size of the study population.

## Ethics approval

This *study* was ethically self-assessed against the ethical principles of the National Statistician’s Data Ethics Advisory Committee (NSDEC) using NSDEC’s ethics self-assessment tool.

## Transparency statement

The manuscript’s guarantor affirms that this manuscript is an honest, accurate, and transparent account of the study being reported; that no important aspects of the study have been omitted; and that any discrepancies from the study as planned (and, if relevant, registered) have been explained.

## Supplementary tables and figures

**Supplementary Table 1.**
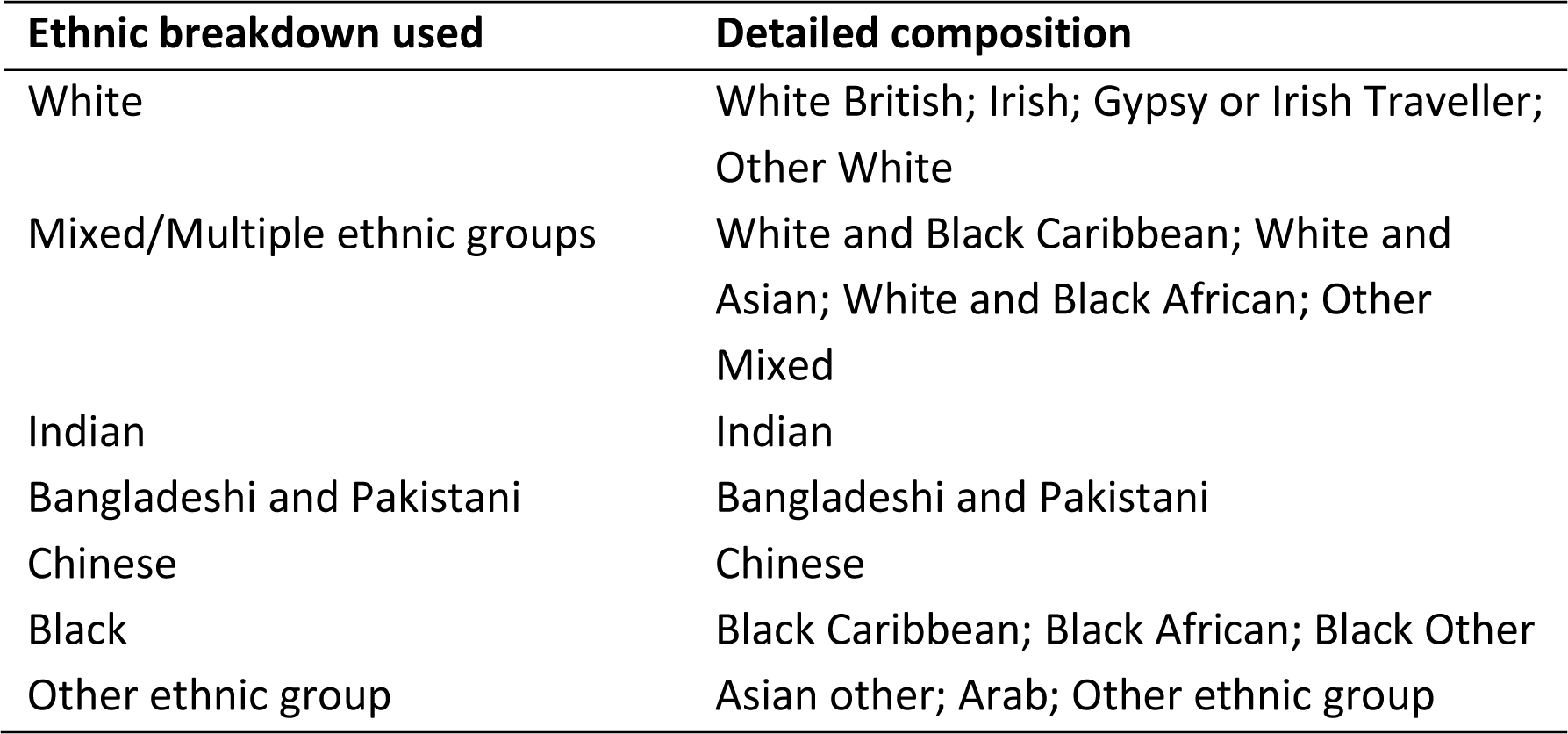
Ethnic breakdowns used in the analysis

| **Ethnic breakdown used** | **Detailed composition** |
| --- | --- |
| White | White British; Irish; Gypsy or Irish Traveller; Other White |
| Mixed/Multiple ethnic groups | White and Black Caribbean; White and Asian; White and Black African; Other Mixed |
| Indian | Indian |
| Bangladeshi and Pakistani | Bangladeshi and Pakistani |
| Chinese | Chinese |
| Black | Black Caribbean; Black African; Black Other |
| Other ethnic group | Asian other; Arab; Other ethnic group |

**Supplementary Table 2.** Age standardised mortality rates (ASMRs) of death involving COVID-19 per 100,000 of the population, stratified by sex and ethnic group

| **Ethnic group** | **Males** | **Females** |
| --- | --- | --- |
| Bangladeshi and Pakistani | 190.4 (172.4, 208.4) | 99.3 (86.6, 112.0) |
| Black | 250.6 (233.2, 268.1) | 116.9 (106.7, 127.2) |
| Chinese | 116.0 (91.1, 145.4) | 64.7 (47.7, 85.6) |
| Indian | 157.9 (145.0, 170.7) | 84.3 (75.5, 93.1) |
| Mixed | 145.2 (120.9, 169.5) | 72.9 (58.5, 89.6) |
| Other | 163.4 (146.1, 180.6) | 80.8 (69.6, 91.9) |
| White | 87.3 (86.0, 88.6) | 51.8 (51.0, 52.7) |

Note: ASMRs are standardised to the study population age distribution. 95% confidence intervals of the ASMRs in parentheses.

**Supplementary Table 3.**
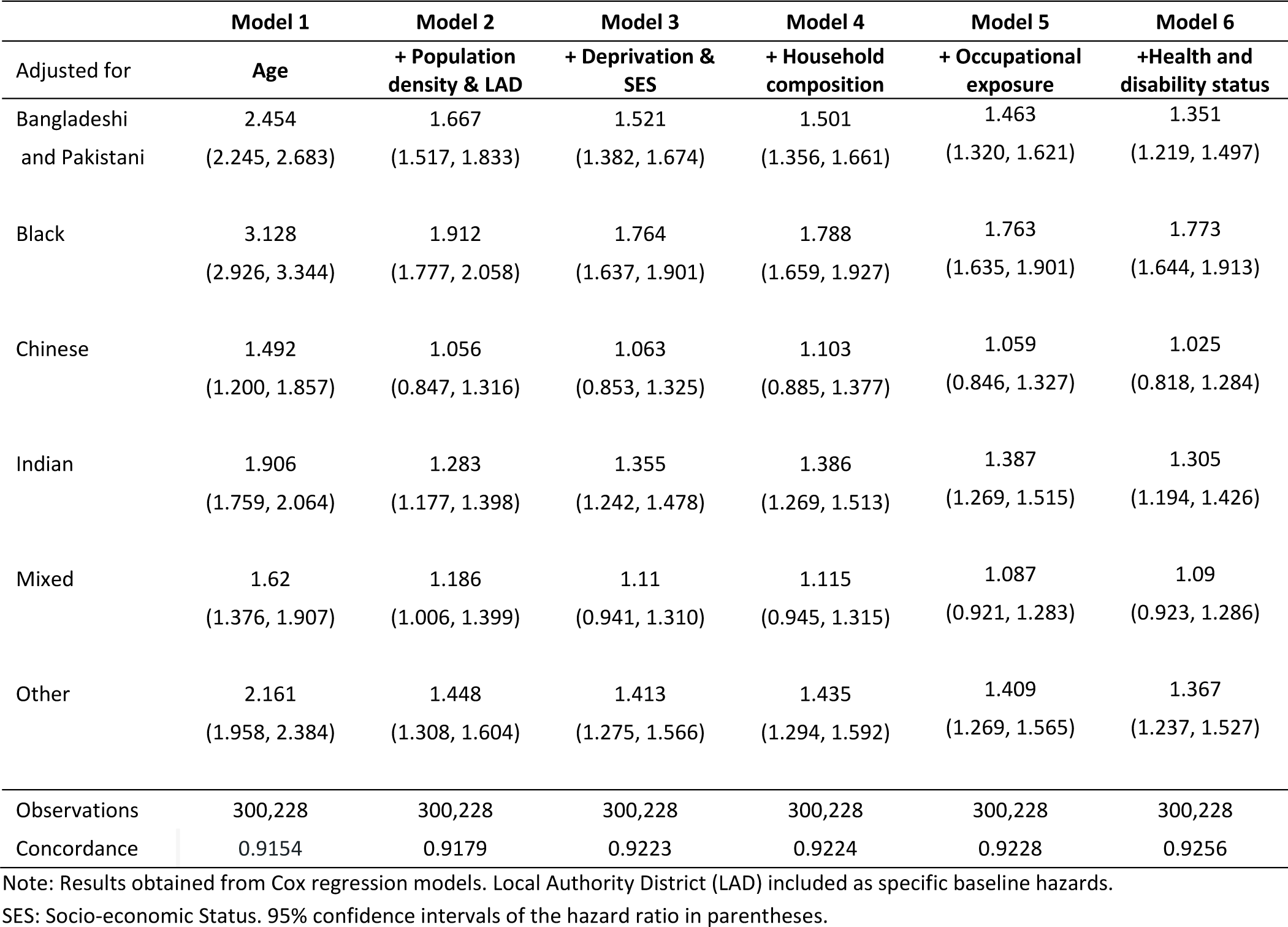
Hazard ratios for COVID-19 related death for ethnic minority groups compared to the White population, adjusted for various covariate sets, males

**Supplementary Table 4.**
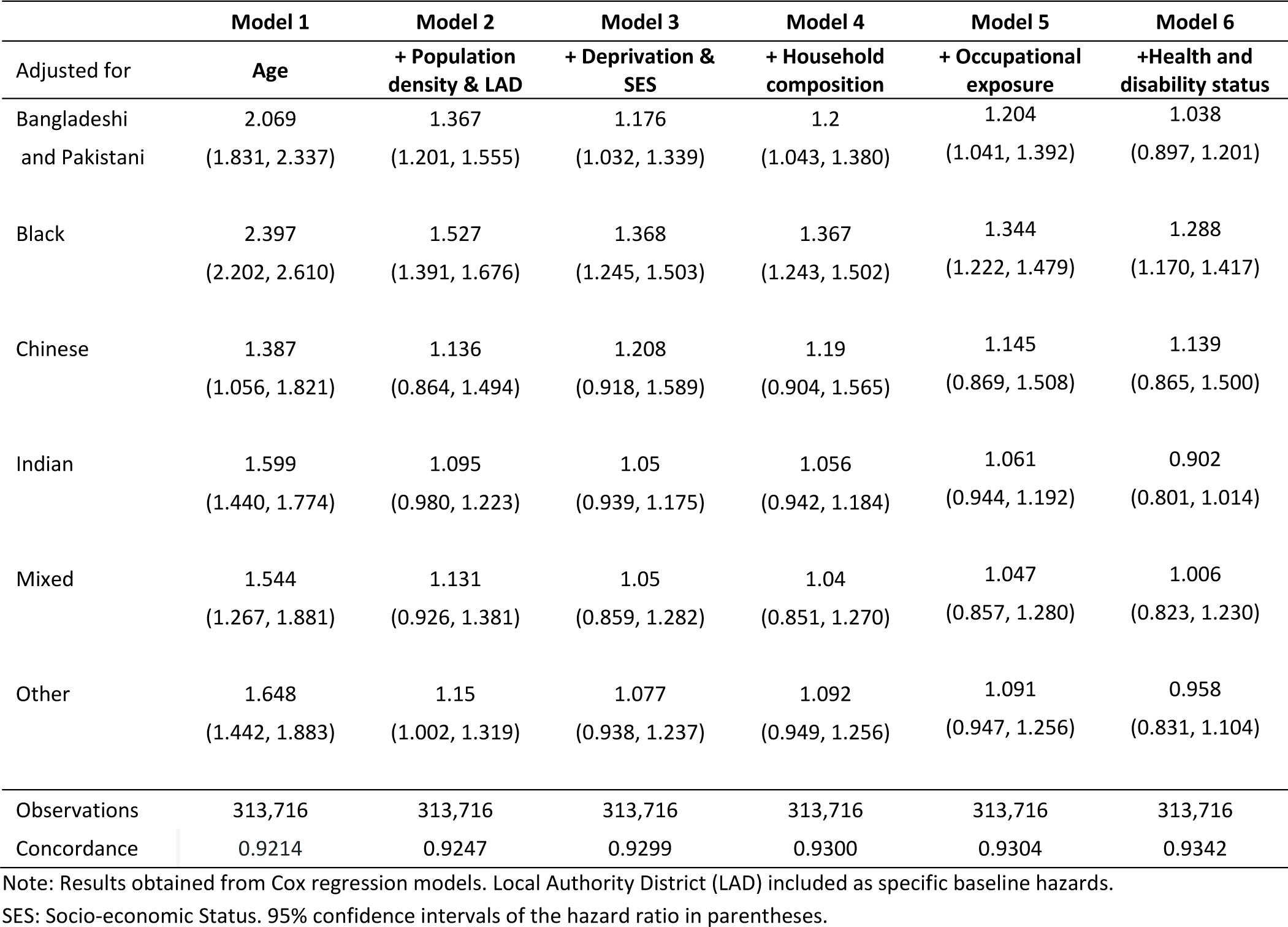
Hazard ratios for COVID-19 related death for ethnic minority groups compared to the White population, adjusted for various covariate sets, females

|  | **Model 1** | **Model 2** | **Model 3** | **Model 4** | **Model 5** | **Model 6** |
| --- | --- | --- | --- | --- | --- | --- |
| Adjusted for | **Age** | **+ Population density & LAD** | **+ Deprivation & SES** | **+ Household composition** | **+ Occupational exposure** | **+Health and disability status** |
| Bangladeshi and Pakistani | 2.069<br>(1.831, 2.337) | 1.367<br>(1.201, 1.555) | 1.176<br>(1.032, 1.339) | 1.2<br>(1.043, 1.380) | 1.204<br>(1.041, 1.392) | 1.038<br>(0.897, 1.201) |
| Black | 2.397<br>(2.202, 2.610) | 1.527<br>(1.391, 1.676) | 1.368<br>(1.245, 1.503) | 1.367<br>(1.243, 1.502) | 1.344<br>(1.222, 1.479) | 1.288<br>(1.170, 1.417) |
| Chinese | 1.387<br>(1.056, 1.821) | 1.136<br>(0.864, 1.494) | 1.208<br>(0.918, 1.589) | 1.19<br>(0.904, 1.565) | 1.145<br>(0.869, 1.508) | 1.139<br>(0.865, 1.500) |
| Indian | 1.599<br>(1.440, 1.774) | 1.095<br>(0.980, 1.223) | 1.05<br>(0.939, 1.175) | 1.056<br>(0.942, 1.184) | 1.061<br>(0.944, 1.192) | 0.902<br>(0.801, 1.014) |
| Mixed | 1.544<br>(1.267, 1.881) | 1.131<br>(0.926, 1.381) | 1.05<br>(0.859, 1.282) | 1.04<br>(0.851, 1.270) | 1.047<br>(0.857, 1.280) | 1.006<br>(0.823, 1.230) |
| Other | 1.648<br>(1.442, 1.883) | 1.15<br>(1.002, 1.319) | 1.077<br>(0.938, 1.237) | 1.092<br>(0.949, 1.256) | 1.091<br>(0.947, 1.256) | 0.958<br>(0.831, 1.104) |
| Observations | 313,716 | 313,716 | 313,716 | 313,716 | 313,716 | 313,716 |
| Concordance | 0.9214 | 0.9247 | 0.9299 | 0.9300 | 0.9304 | 0.9342 |

Note: Results obtained from Cox regression models. Local Authority District (LAD) included as specific baseline hazards. SES: Socio-economic Status. 95% confidence intervals of the hazard ratio in parentheses.

**Supplementary Table 5.** Age standardised mortality rates (ASMRs) of death involving COVID-19 per 100,000 of the population, before and after lockdown plus 21 days, stratified by sex and ethnic group

| Ethnic group | Males |  | Females |  |
| --- | --- | --- | --- | --- |
|  | Pre-lockdown | Post-lockdown | Pre-lockdown | Post-lockdown |
| Bangladeshi and Pakistani | 116.9 (102.8, 131.0) | 74.2 (62.9, 85.6) | 54.8 (45.4, 64.3) | 44.8 (36.2, 53.3) |
| Black | 157.0 (143.1, 171.0) | 95.2 (84.5, 106.0) | 79.9 (71.4, 88.4) | 37.5 (31.7, 43.3) |
| Chinese | 75.7 (55.8, 100.2) | 40.9 (26.7, 59.7) | 30.0 (19.0, 44.8) | 35.0 (22.7, 51.4) |
| Indian | 87.9 (78.3, 97.5) | 70.8 (62.1, 79.4) | 47.2 (40.6, 53.8) | 37.5 (31.6, 43.4) |
| Mixed | 74.5 (58.1, 94.1) | 71.6 (55.4, 90.9) | 34.5 (24.9, 46.4) | 38.7 (28.4, 51.5) |
| Other | 95.0 (81.8, 108.2) | 69.1 (57.9, 80.3) | 42.0 (34.0, 50.0) | 39.0 (31.2, 46.9) |
| White | 38.0 (37.1, 38.8) | 50.1 (49.1, 51.1) | 19.5 (18.9, 20.0) | 32.8 (32.1, 33.4) |

Note: ASMRs are standardised to the study population age distribution. 95% confidence intervals of the ASMRs in parentheses.

**Supplementary Figure 1.**
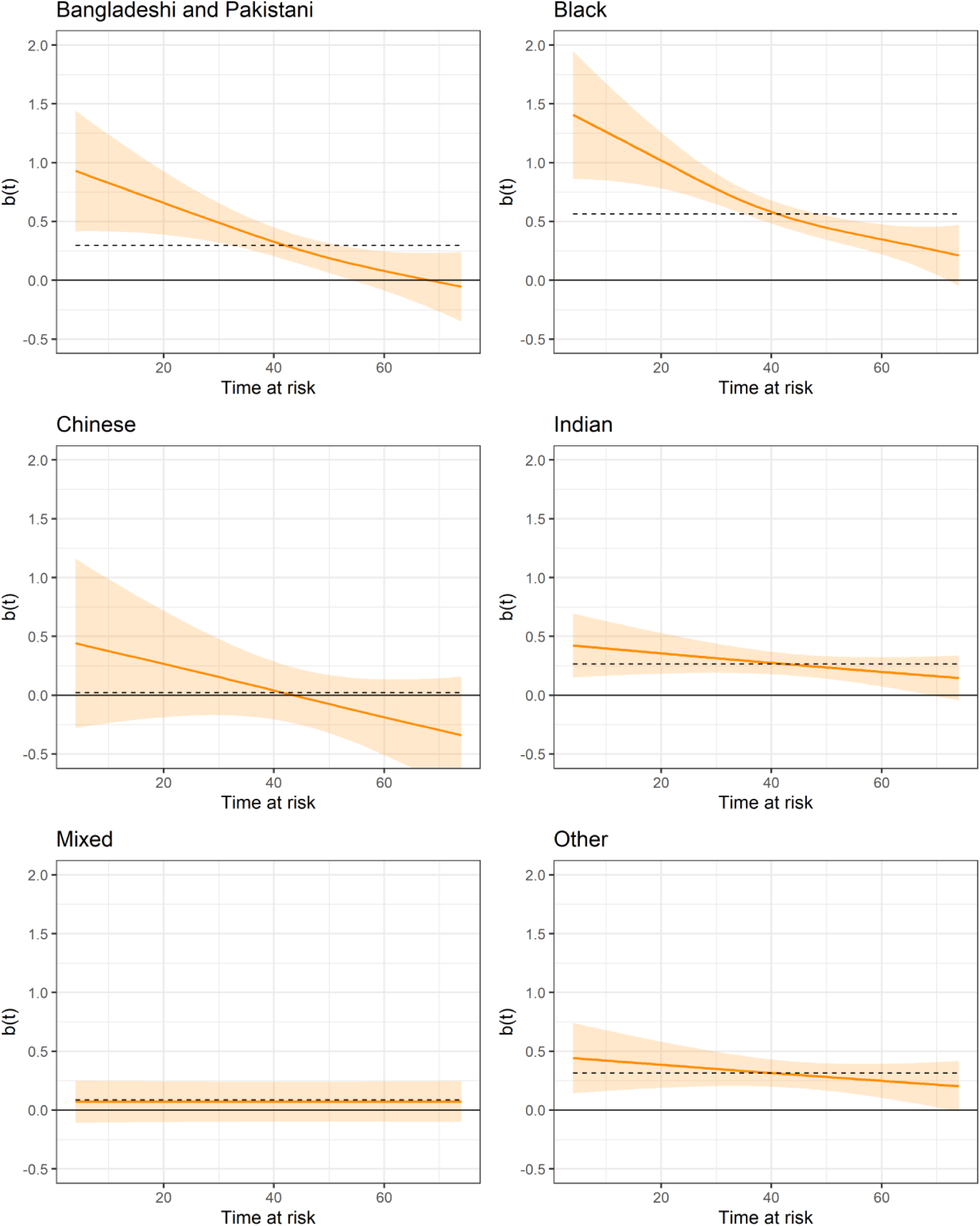
Smoothed Schoenfeld residuals for ethnic minority groups from the fully adusted model, males Note: Results obtained from a Cox regression model adjusted for age, population density, deprivation, SES, household composition, occupational exposure, self-reported health, with baseline hazards specific to local authority district. Residuals are smoothed with a generalised additive model. Confidence intervals are at the 95% level.

**Supplementary Figure 2.**
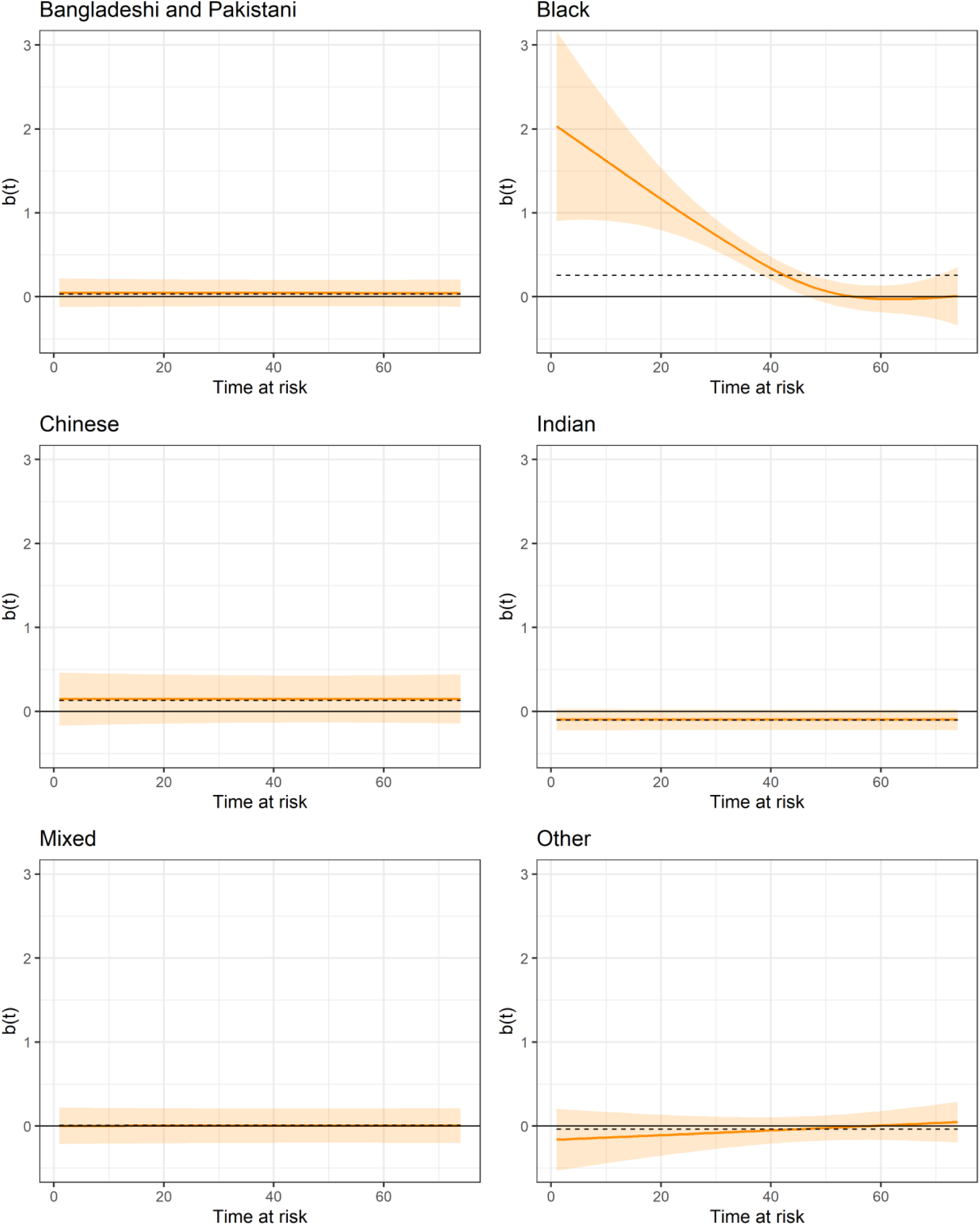
Smoothed Schoenfeld residuals for ethnic minority groups from the fully adusted model, females Note: Results obtained from a Cox regression model adjusted for age, population density, deprivation, SES, household composition, occupational exposure, self-reported health, with baseline hazards specific to local authority district. Residuals are smoothed with a generalised additive model. Confidence intervals are at the 95% level.

**Supplementary Figure 3.**
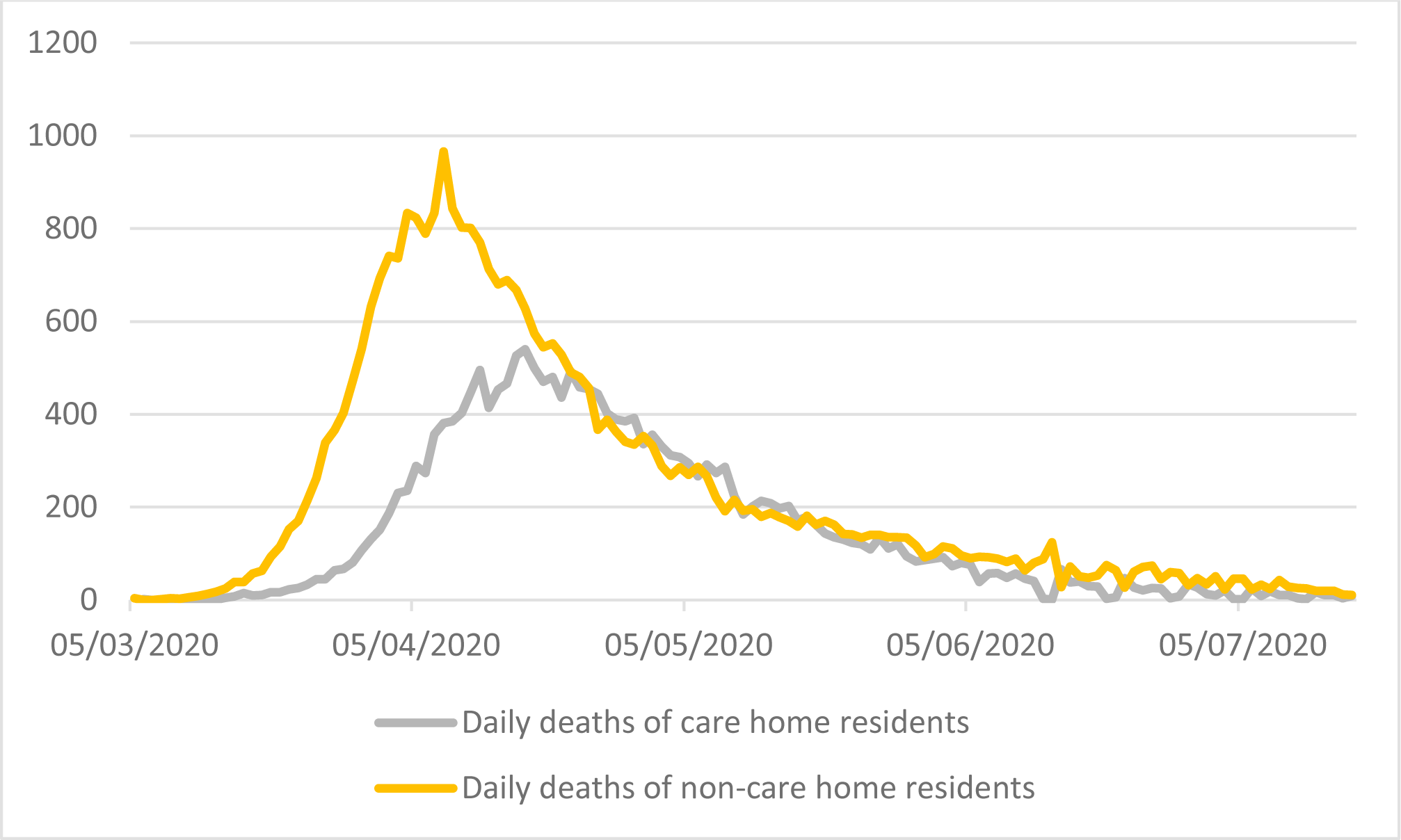
Daily deaths in care homes and in the community Data: ONS care home deaths; ONS daily deaths

**Supplementary Figure 4.**
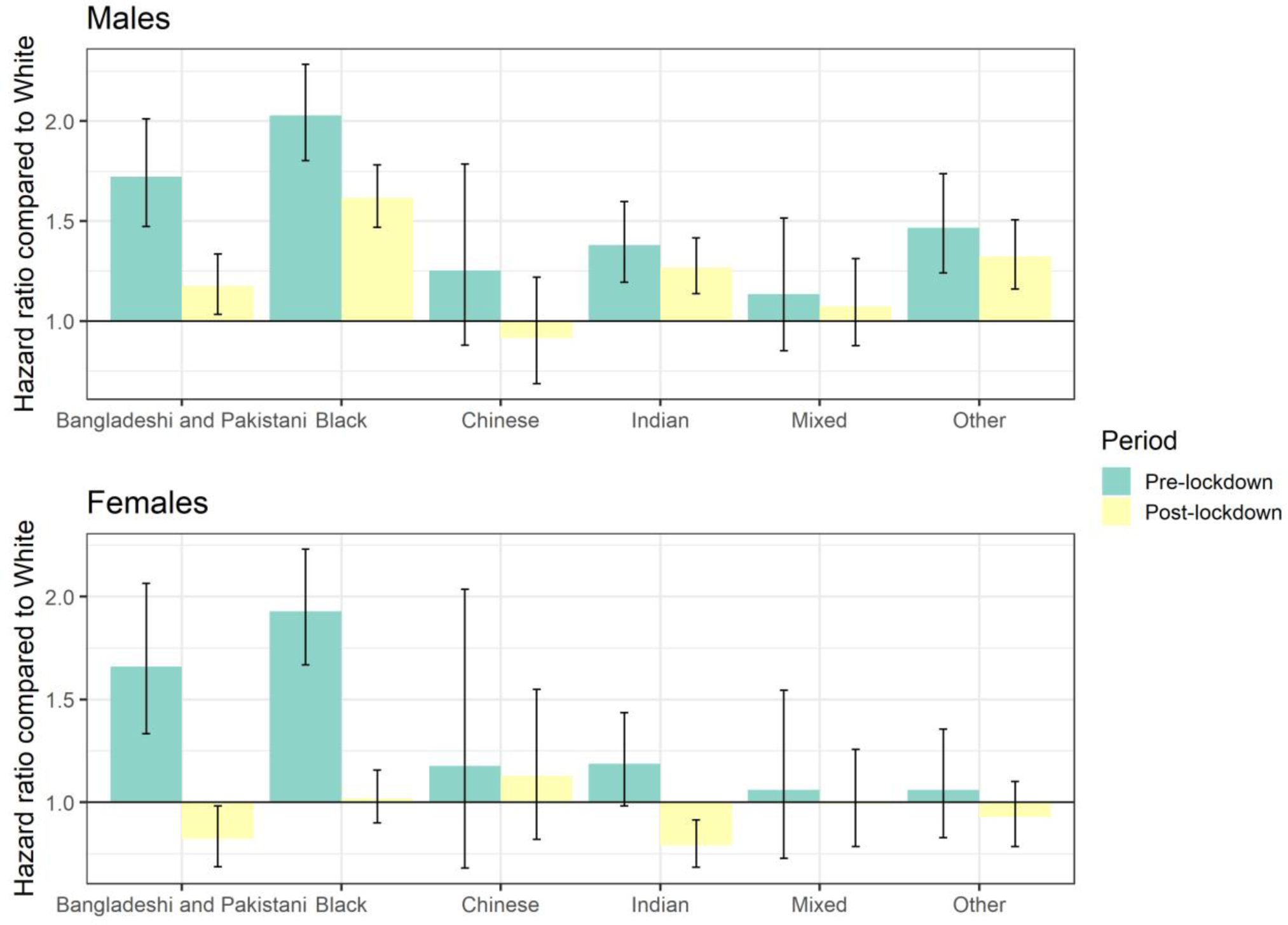
Hazard ratios for COVID-19 related death for ethnic minority groups compared to the White population, before and after lockdown plus 14 days, stratified by sex

**Supplementary Figure 5.**
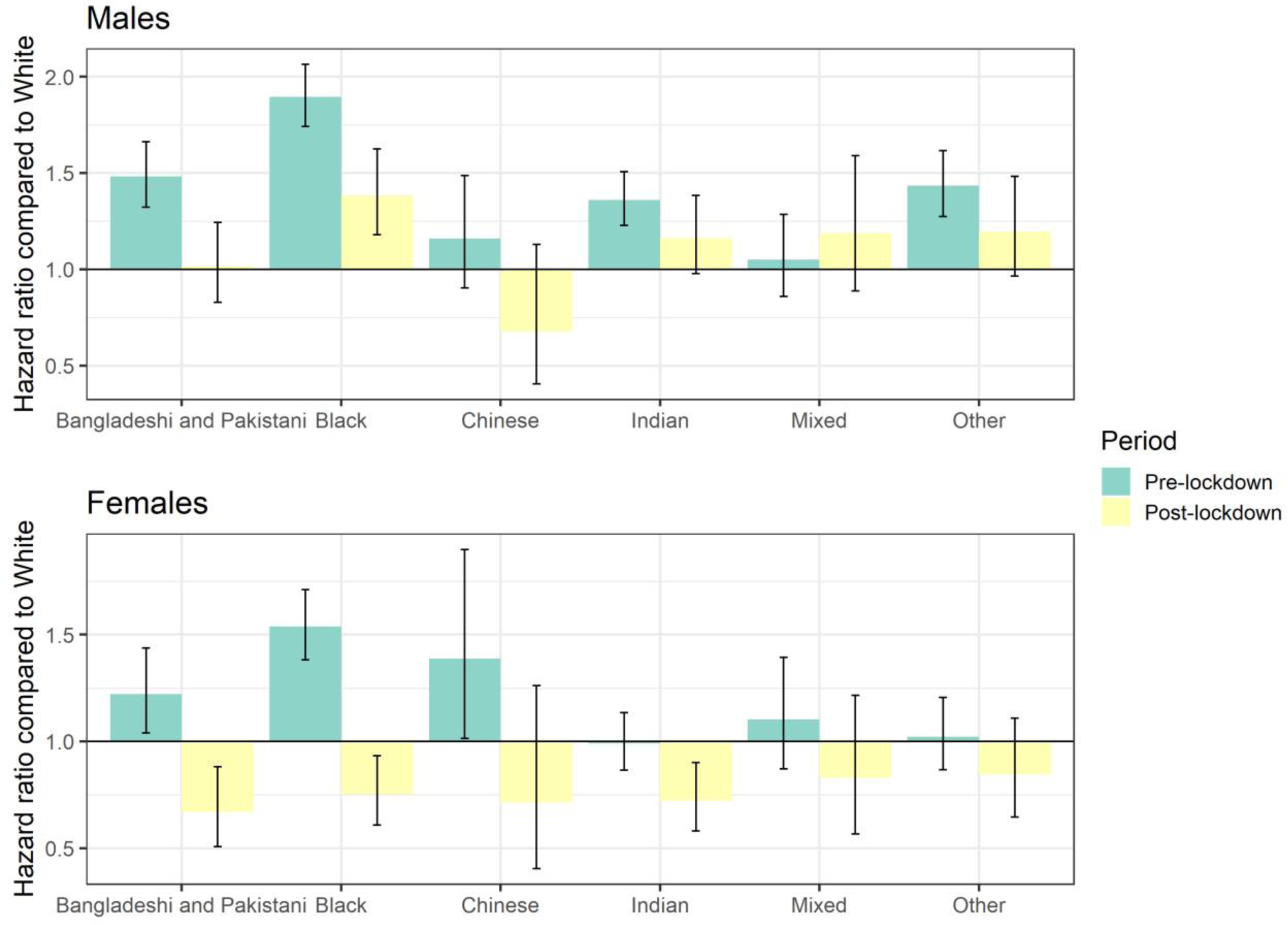
Hazard ratios for COVID-19 related death for ethnic minority groups compared to the White population, before and after lockdown plus 28 days, stratified by sex

## Supplementary appendix on data linkage

### Linkage methodology

The 2011 Census was linked to the 2011-2013 Patient Registers using deterministic and probabilistic matching. It was first linked deterministically using 24 different matching keys, based on a combination of forename, surname, date of birth, sex and geography (postcode or Unique Property Reference Number). Using different combinations of these variables ensured that records that contain errors in these variables could nonetheless be linked. The matches must have been unique within a matching key for the match to be accepted.

Probabilistic matching was then used to attempt to match records that were not linked deterministically, using 13 different combinations of personal identifiers. Candidate matches were assigned to Census records using the Felligi-Sunter probabilistic matching method.

Of the 53,483,502 Census records, 50,019,451 were linked deterministically. 555,291 additional matches were obtained using probabilistic matching.

### Linkage rate by characteristics

Table 3 shows the proportion of Census records that were linked to the Patient Register for different groups. These linkage rates are based on the entire 2011 Census population.

**Table 3.**
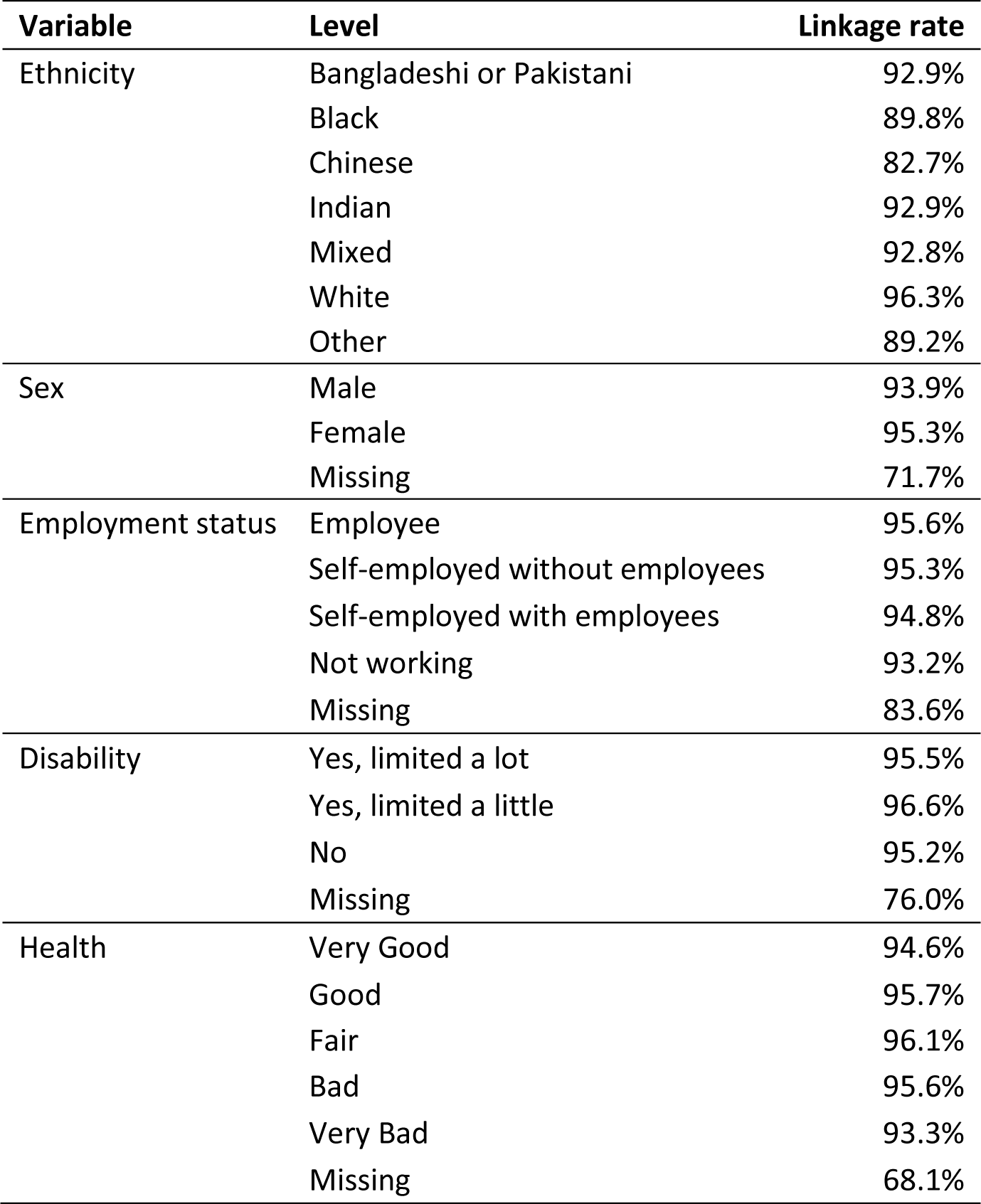
Linkage rates between the 2011 Census and the 2011-2013 Patient Registers by selected individual characteristics

| Variable | Level | Linkage rate |
| --- | --- | --- |
| Ethnicity | Bangladeshi or Pakistani | 92.9% |
|  | Black | 89.8% |
|  | Chinese | 82.7% |
|  | Indian | 92.9% |
|  | Mixed | 92.8% |
|  | White | 96.3% |
|  | Other | 89.2% |
| Sex | Male | 93.9% |
|  | Female | 95.3% |
|  | Missing | 71.7% |
| Employment status | Employee | 95.6% |
|  | Self-employed without employees | 95.3% |
|  | Self-employed with employees | 94.8% |
|  | Not working | 93.2% |
|  | Missing | 83.6% |
| Disability | Yes, limited a lot | 95.5% |
|  | Yes, limited a little | 96.6% |
|  | No | 95.2% |
|  | Missing | 76.0% |
| Health | Very Good | 94.6% |
|  | Good | 95.7% |
|  | Fair | 96.1% |
|  | Bad | 95.6% |
|  | Very Bad | 93.3% |
|  | Missing | 68.1% |

The linkage rates exceeded 80% for all ethnic groups. The White ethnic group had the highest linkage rate (96.3%) whilst those from Chinese background had the lowest (82.7%). To an extent, these differences in linkage rates reflect differences in the age distributions of the ethnic groups. As shown in Figure 1, the linkage rate was lower for young adults than for children or older adults; however, the linkage rate was still above 84% for every age group.

**Figure 1.**
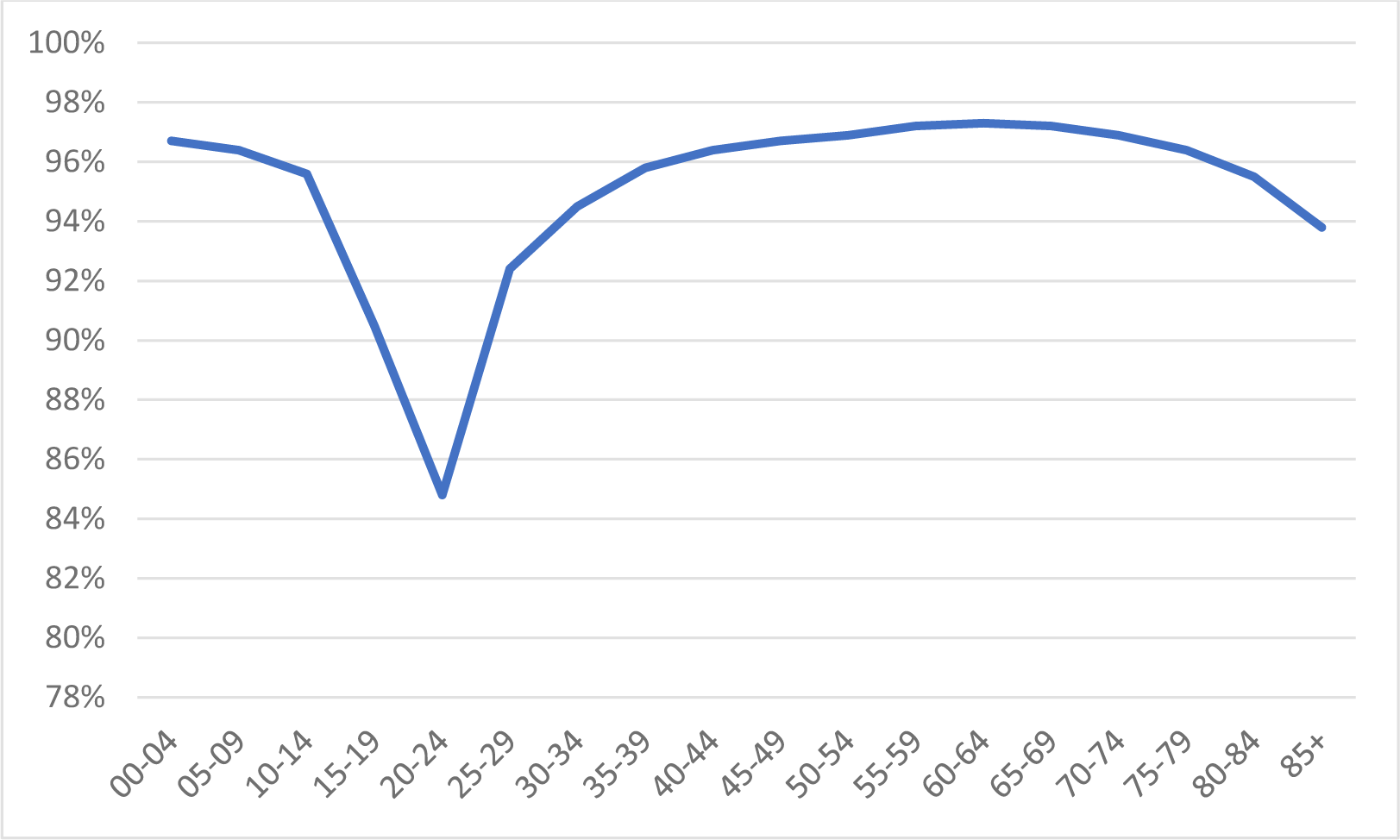
Linkage rates between the 2011 Census and the 2011-2013 Patient Registers by age

The linkage rate was slightly higher for females (95.3%) than males (93.9%). There was little difference in linkage rate by employment status, although the rate was slightly lower among those who were not working. This could be in part driven by age, as those of student age tended to have a lower linkage rate.

The linkage rate was very similar across disability groups: 95.2% for those who did not report any disability, compared to 96.6% and 95.5% for those who reported having their daily activity limited a little or a lot, respectively. The linkage rate did not vary substantially depending on self-reported health status.

Across all variables, the linkage rate was high for all groups other than those reporting a missing or invalid code. The linked population can therefore be said to be broadly representative of the general population of England and Wales in 2011.

## Supplementary appendix on adjusting for emigration

### Introduction

Embarkation weights based on survey and administrative data accounted for emigration from the study population between the 2011 Census and 2 March 2020. Adjusting for emigrants (as well as deaths) over this period allowed us to re-weight our analysis to the study population at risk of death from 2 March 2020.

The embarkation weights were based on observed and unobserved embarkation information from the NHS Patient Register linked to the ONS Longitudinal Study (LS), and on measures derived from the International Passenger Survey (IPS) and applied to the linked Census-Patient Register-Deaths dataset.

To adjust for emigration, we estimated the probability to emigrate in every year between March 2011 and March 2020 by sex, age group and ethnicity using data from the NHS Patient Register and the IPS. The probability of remaining in the country was calculated as the complement of the probability of emigrating, and from this we obtained inverse probability weights.

IPS data were only available up to year ending March 2019, we assumed emigration rates observed between March 2019 and March 2020 were the same as those observed in the previous year. We then combined these annual probabilities to create a weight indicating each individual’s probability of remaining in the country between March 2011 and March 2020. We applied these migration adjustment factors to all analyses presented in the paper.

### Rates based on the ONS Longitudinal Study for England and Wales

We calculated embarkation rates for a study cohort using NHS Patient Register data linked to the LS for England and Wales for 2011-2016. The LS cohort is a 1% sample of those present at the 2011 Census. These rates were adjusted for mortality for usual residents only, by sex, age group and ethnicity.

Embarkations were identified in the LS using NHS Patient Register data, where a person deregistered with a GP because they had or were moving abroad (observed embark), or where a patient registration was cancelled by the GP surgery (unobserved embark).

### Rates based on the International Passenger Survey

We reviewed all available data sources to calculate comparative embarkation rates, including the IPS and Annual Population Survey (APS). We concluded that the IPS was the best available contemporaneous data source with estimates available up to year ending March 2019.

The IPS does not collect data on ethnicity but does collect data on citizenship. We therefore produced an IPS based out-migration rate via a lookup between citizenship and ethnic group by age and sex, using published 2011 Census tables for England and Wales.

We equivalised the IPS data to mimic a cohort study. To do this we excluded anyone who immigrated after the 2011 Census and subsequently emigrated in the time period of interest (2011-2020). We adjusted IPS emigrants in age groups 0-4 years and 5-9 years to mimic a cohort with non-replacement, using a uniform distribution. For example, we decreased the number of emigrants in the 0-4 age group by 1/5 in 2012/13, reflecting non-replenishment in the cohort. We decreased this age group by 1/4 in 2013/14, and so on. This resulted in no one in the 0-4 years age group by 2016/17. In 2017/18, we started to decrease the number of emigrants in the 5-9 age group.

We took the Census microdata as our starting population in 2011/12 and only included usual residents. To move to our new base population in 2012/13, we subtracted those who emigrated in 2011/12 from the base population by age group and ethnicity. We then deducted deaths by age group and ethnicity.

We aged the 2012/13 base population on a year. We assumed a constant age distribution whereby we aged-on 1/25 of those aged 0-24 years and added into ages 25-44 years, for 25-44 we aged-on 1/20 of the group and added into ages 45-64 years, and so on.

We also needed to account for not adding in new-borns over time, reflecting the fact that the 0-24 years age group would have become 1-24 years in 2012/13, 2-24 years in 2013/14, and so on.

Embarkation rates for the cohort by sex, age group (0-24 years, 25-44 years, 45-64 years, 65+ years), ethnicity (White, Mixed, Indian, Chinese, Pakistani and Bangladeshi, Black, Other) and year (2011/12 through to 2019/20) were calculated using the following formula:

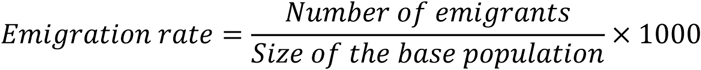

### Extrapolation and hybrid approach

We developed a hybrid approach that built on the strength of the relationship between the LS-(usual resident-based) and IPS-based rates for 2012/13 through to 2015/16. This addressed the shortcoming of LS-based rates only being available up to 2015/16 and IPS-based rates to 2018/19.

The methodology took the mean of LS- and IPS-based rates for years 2012/13 through to 2015/16 and extended this for the rest of the nine-year period.

- For 2011/12, the ratio of LS to LS-IPS mean rates in 2012/13 was applied to LS rates in 2011/12 to derive a new mean for 2012/13.
- For 2016/17 through to 2019/20, the absolute difference between the IPS-LS mean and the IPS rate in 2015-16 was applied to IPS rates for 2016/17, 2017/18 and 2018/19 to extrapolate new means for each group in these years. The 2018/19 rates were repeated for 2019/20.

